# Portable Cerebral Blood Flow Monitor to Detect Large Vessel Occlusion in Suspected Stroke Patients

**DOI:** 10.1101/2023.12.14.23299992

**Authors:** Christopher G. Favilla, Grayson L. Baird, Kedar Grama, Soren Konecky, Sarah Carter, Wendy Smith, Rebecca Gitlevich, Alexa Lebron-Cruz, Arjun G. Yodh, Ryan A. McTaggert

**Author notes:** Corresponding author: Christopher Favilla, 3400 Spruce St, 3 West Gates Bldg, Department of Neurology, Philadelphia, PA 19104. These two authors contributed eqaully to the manuscript (Christopher G. Favilla, Grayson L. Baird).

## Abstract

**Background:** Early detection of large vessel occlusion (LVO) facilitates triage to a comprehensive or thrombectomy-capable stroke center to reduce treatment times and improve outcomes. Prehospital stroke scales, however, are not sufficiently sensitive, so here we investigate the ability of the portable Openwater optical blood flow monitor to detect LVO in patients undergoing acute stroke evaluation.

**Methods:** Patients were prospectively enrolled at two comprehensive stroke centers during acute stroke evaluation within 24 hours of symptom onset with NIHSS ≥2. Each patient underwent a 60-second bedside optical blood flow evaluation with the Openwater instrument. The Openwater instrument generates cerebral blood flow waveforms based on relative changes in speckle contrast. Anterior circulation LVO was deteremined based on CT angiographic imaging and defined as occlusion of the ICA, or first/second segment of the MCA. A deep learning model, based on a transformer architecture trained on all patient data using fivefold cross-validation and learned discriminative representations from the raw speckle contrast waveform data. ROC analysis compared the Openwater diagnostic performance (i.e., LVO detection) with performance of prehospital stroke scales.

**Results:** Amongst 135 patients, the median NIHSS was 8 (IQR: 4-14), and 52 (39%) had an anterior circulation LVO based on CT angiogram. The Openwater instrument had 79% sensitivity and 84% specificity for the detection of LVO. The RACE scale had 60% sensitivity and 81% specificity. LAMS had 50% sensitivity and 81% specificity. In the ROC analysis, the binary Openwater classification (high-likelihood vs low-likelihood) had an AUC of 0.82 (95%CI: 0.75-0.88), which outperformed RACE (AUC: 0.70; 95%CI: 0.62-0.78; p=0.04) and LAMS (AUC: 0.65; 95% CI: 0.57-0.73; p=0.002).

**Conclusions:** The Openwater optical blood flow monitor outperformed prehospital stroke scales for the detection of LVO in patients who presented to the Emergency Department for acute stroke evaluation. These encouraging findings need to be validated in the prehospital environment.

## INTRODUCTION

Endovascular therapy (EVT) has revolutionized treatment of acute stroke with large vessel occlusion (LVO)^1,2^ but is only available at a minority of stroke centers.^3,4^ Early LVO recognition during pre-hospital care presents an opportunity to route patients to endovascular capable centers and thereby reduce treatment times and improve outcomes.^5–8^ In fact, the American Heart Association along with its Mission: Lifeline® Stroke algorithm recommend that emergency health services (EMS) route high-likelihood LVO patients to comprehensive or thrombectomy-capable stroke centers, depending on the additional transportation time.^9,10^ In current practice, the likelihood of LVO is most often determined by one of several pre-hospital stroke severity scales for which diagnostic accuracy is suboptimal, and implementation in clinical practice is inconsistent.^11–15^ Thus, several non-invasive portable technologies have been explored with a goal to develop a wearable LVO detector.^16,17^ Transcranial Doppler, volumetric impedance phase shift spectroscopy (VIPS), and electroencephalography (EEG) have been studied to this end with varying degrees of diagnostic accuracy.^18–21^ Lastly, in addition to performance metrics, it is important to consider cost, size, efficiency, and ease of use in the pre-hospital setting.^22^

A direct cerebral blood flow (CBF) monitor is an intuitive choice for development of LVO detectors. TCD-based CBF waveform morphology is reasonably sensitive and specific for LVO,^23,24^ but the technique is limited by time, cost, and the need for technical expertise.^25^ A robotic TCD may resolve the need for technical expertise^26^ but at higher unit cost; furthermore, nearly 20% of the population does not have adequate temporal acoustic windows.^27^ Biomedical optical imaging (*i.e.,* diffuse optical monitoring) offers a promising alternative for directly probing tissue-level physiology,^28,29^ and prior work has demonstrated the ability for monitoring acute stroke physiology at the bedside.^30,31^ The Openwater optical blood flow monitor (Openwater; San Francisco, CA) is a novel, wearable device that leverages measurements of speckle contrast and light intensity to continuously monitor microvascular hemodynamics and resolve a pulsatile CBF waveform in a cost-effective portable instrument.^32^ Here, we evaluate the diagnostic performance of the Openwater optical blood flow monitor to detect the presence of LVO in patients presenting for an acute stroke evaluation.

## METHODS

### Participants

Eligible patients were prospectively enrolled in this observational cohort at two comprehensive stroke centers (Hospital of the University of Pennsylvania and Rhode Island Hospital) if they presented to the emergency department or were transferred from another facility for acute stroke management within 24 hours of symptom onset and underwent emergent neurovascular imaging as per routine care to evaluate for possible LVO. Eligible patients had National Institutes of Health Stroke Scale (NIHSS) ≥ 2. Patients were excluded if they had a known intracranial mass, a skull defect that would interfere with optical monitoring, or clinical suspicion for bilateral infarcts. Patients were enrolled between August 22, 2022 and May 30, 2023.

The study was approved by the University of Pennsylvania and Lifespan Institutional Review Boards, and informed consent was provided by each subject (or legally authorized surrogate).

### Clinical and neuroimaging data

Patient demographics and baseline characteristics, stroke timing, presenting NIHSS, rapid arterial occlusion evaluation (RACE) scale, Los Angeles motor scale (LAMS), were extracted from the electronic health record. Given the potential relevance of skin pigmentation to optical data quality, the Fitzpatrick scale was used categorize skin color for each patient. Baseline CT results were reviewed to confirm the presence or absence of intracerebral hemorrhage. Baseline CT angiogram results were reviewed to confirm the presence or absence of LVO. For study purposes, LVO was defined as occlusion of the cervical or intracranial internal carotid artery (ICA), M1 segment of the MCA, M2 segment of the MCA, or tandem occlusion. For non-LVO patients, the electronic health record was reviewed at discharge to confirm the final diagnosis, and non-LVO patients were further categorized as (1) ischemic stroke without LVO, (2) intracerebral hemorrhage, or (3) stroke mimic (mimics were further sub-categorized as seizure, migraine, conversion disorder, or other).

### Openwater optical blood flow monitor

The hemodynamic measurement device (Openwater; San Francisco, CA) consists of a wearable headset and a console (**Figure 1**). The headset contains two modules that collect data simultaneously from both sides of the head. The headset size was adjustable via a built-in dial. Each module contains a built-in optical fiber for the delivery of low average power laser light to the surface of the brain, and each light source is associated with three custom cameras for the measurement of light escaping from the subject. The console contains the laser, electronics, touchscreen, and computer. The optical methodology was previously described in detail.^32^

**Figure 1.**
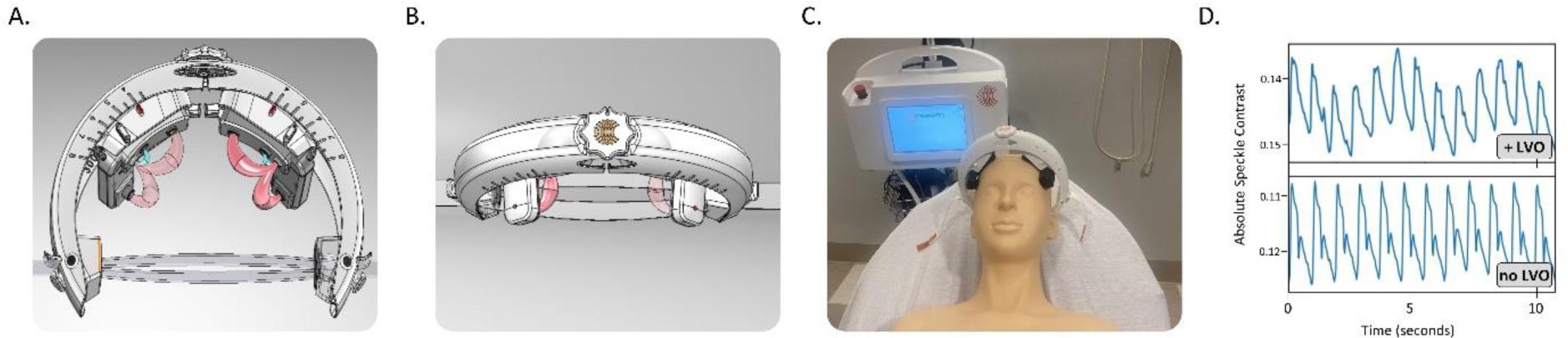
Instrumentation and waveform data: (A) A schematic of the Openwater headset depicts the light source/detector positioning and the theoretical light path, and (B) an additional schematic depicts an anterior view. (C) A photograph depicts the Openwater headset positioning on the head with the console in the background. (D) The speckle contrast derived CBF waveforms are depicted from two representative subjects (one with LVO and one without). CBF indicates cerebral blood flow. LVO indicates large vessel occlusion.

### Optical CBF evaluation

Each patient underwent a 70-second bedside optical blood flow evaluation with the Openwater system after presenting to the comprehensive stroke center for acute stroke evaluation. All evaluations were performed within 24 hours of symptoms onset. If the exact onset was unknown, the time last known well was used as a surrogate for onset time. For LVO patients, the CBF evaluation was completed prior to EVT (if applicable). With the patient lying supine in the flat hospital bed or stretcher, the Openwater headset was placed on the patient’s head and positioned such that the optical probes were at the superior aspect of the forehead. The built-in dial was adjusted to position the dial at the lateral margin of the forehead (while avoiding hair). The elastic strap was then tightened to ensure adequate contact between the skin and optical probes. A 70-second scan was performed across the six detectors. The speckle contrast-derived CBF waveform was acquired at 40 Hz. Representative waveforms were depicted in **Figure 1d**.

After data acquisition, two data quality checks were performed. First, ambient light and laser light levels were assessed to ensure probe contact was appropriate throughout the scan. The scan was considered a technical failure if more than 2 (of 6) cameras failed this quality check. Next, the frequency spectrum of data from each sensor were analyzed, and if a consistent pulse was not detected across four or more detectors, the pulse check was considered a failure, and data were rejected.

### LVO detection model

For classification, we used a deep learning model based on a transformer architecture that learns discriminative representations from the raw speckle contrast waveform data. The network was trained on all patient data using fivefold cross-validation.

For classification, we utilized a previously described deep learning model that effectively recognizes ECG waveform abnormalities.^33^ This model employs a transformer architecture, which effectively extracts distinctive feature representations from the speckle contrast waveform data. We leverage self-attention pooling on the outputs of the transformer layers to enhance the model’s performance.^34^ The network’s output is then converted into a probability score for either the LVO or the non-LVO class using the SoftMax function.^35^ To mitigate the data scarcity issue, we implement fivefold cross-validation,^36^ randomly dividing the data into five testing sets. Five independent models are trained, and the reported results are based on the performance on these five independent testing folds using patients that were not included in the corresponding fold during training.

### Design and Statistical analysis

To better generalize study results to the pre-hospital setting, LVO patients were compraed with non-LVO patients who underwent the same acute stroke evaluation (rather than comparing healthy controls). Specifically, to maximize sensitivity and specificity estimation, an enriched sample was collected, where LVO accounted for 38% (51/135) of cases and non-LVO cases accounted for 62% (84/135) of the cohort (non-LVO ischemic stroke, hemorrhage, or stroke mimic). As a reference standard for diagnostic performance, RACE and LAMS were collected and used with their established thresholds ≥5 and ≥4, respectively. No power analysis was conducted since this work was a pilot study to determine initial performance of the Openwater device; data observed here will inform future analyses.

Diagnostic performance was examined using receiver operator characteristic curve area under the curve (AUROC) with the LOGISTIC procedure. Youden’s J was estimated for the Openwater device using the %ROCPLOT macro and was used to determine the mathematically optimal performance of sensitivity and specificity; prediction score, sensitivity, and specificity were estimated using generalized linear model assuming a binary distribution with the GLIMMIX procedure. Positive (PPV) and negative predictive values (NPV) were derived from the sensitivity and specificity estimates along with a prevalence of 5% and 10%, respectively, using Bayes theorem. Alpha was set at the 0.05 level, and all interval estimates were estimated for 95% confidence. All analyses were conducted using SAS 9.4. The data and code that support the reported findings are available from the corresponding author upon reasonable request.

## RESULTS

A total of 162 patients underwent an optical CBF evaluation as part of Emergency Department stroke alert workflow, 135 of whom yielded a fully analyzable sample. **Figure 2** summarizes patient enrollment and network classification. The trained neural network categorized 40% of patients as high likelihood LVO and 60% as low likelihood LVO. Based on clinical vascular imaging, 52 patients (39%) were ultimately found to have a LVO. LVOs consisted of 18% ICA, 40% M1, 24% M2, 18% tandem ICA/MCA. **Table 1** summarizes the cohort demographics, baseline characteristics, and final diagnosis. Patients who were excluded based on a failed optical CBF scan were similar to patients who were included in the final cohort (**Table S1**), but excluded subjects suffered more severe strokes [NIHSS: 19 (8 – 22) versus 8 (4 – 14), p = 0.002] and were more likely to have LVO (67% vs 38%, p = 0.006).

**Figure 2.**
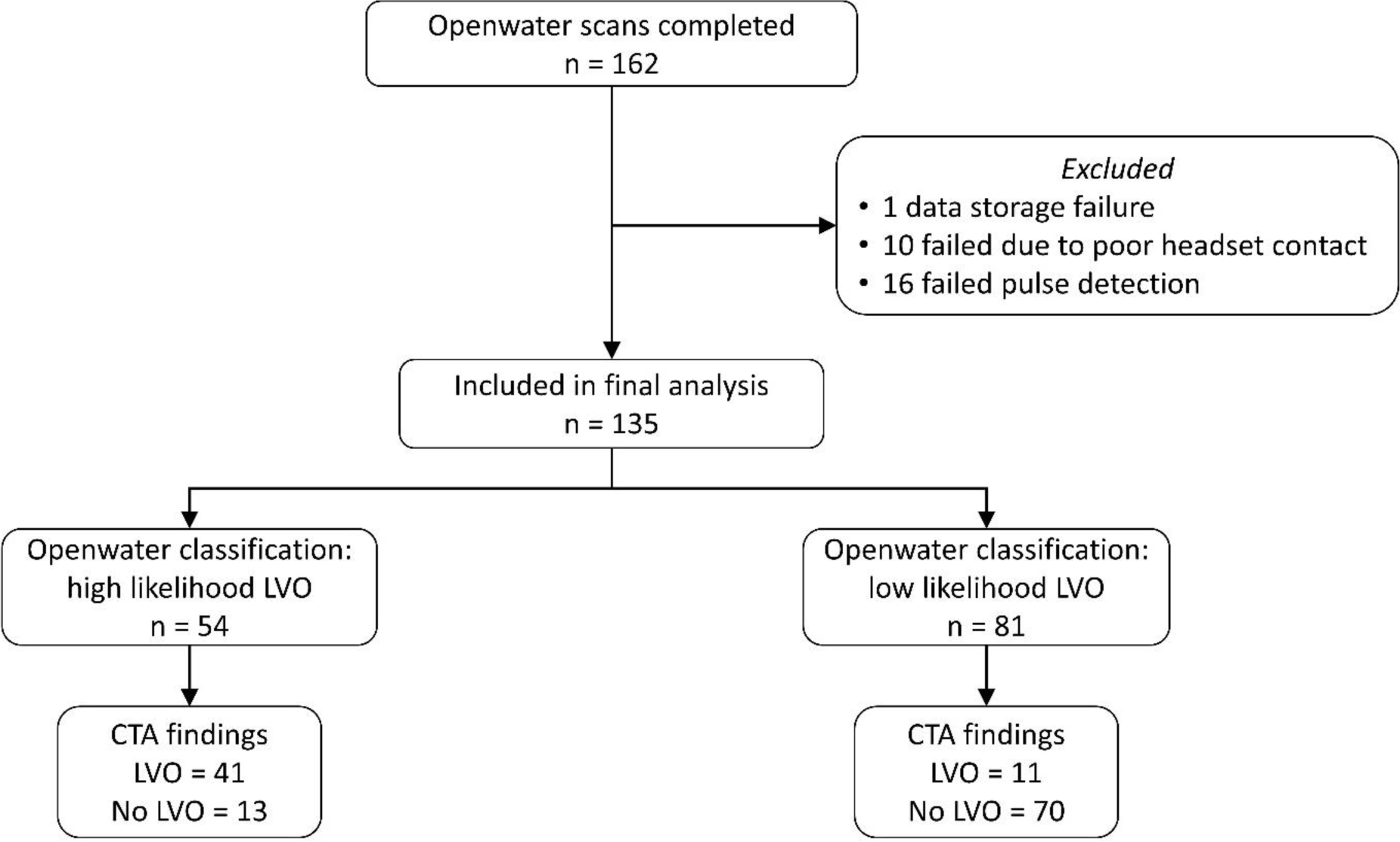
Cohort flow diagram: The Openwater scan was performed in 162 patients. After excluding patients with technically limited scans, 135 were included in the final analysis. The Openwater optical blood flow monitor classified 54 (40%) as high likelihood LVO and 81 (60%) as low likelihood LVO. Based on standard of care CTA, 52 patients were diagnosed with LVO. CTA indicates computer tomography angiography. LVO indicates large vessel occlusion.

**Table 1.** Cohort characteristics.

|  |  |
| --- | --- |
|  | Final cohort<br>n = 135 |
| Age, years | 70 (15) |
| Sex, % female | 46% |
| Race, % |  |
| Asian | 1% |
| Black or African American | 28% |
| White | 66% |
| Unknown | 5% |
| Ethnicity, % Hispanic or Latino | 2% |
| Fitzpatrick Scale | 2 (2 – 4) |
| NIHSS | 8 (4 – 14) |
| Received IV thrombolysis, % | 24% |
| Time from onset to Openwater scan, hours | 7.8 (3.0 – 14.9) |
| Diagnostic categorization, % |  |
| Large vessel occlusion | 39% |
| Ischemic stroke without occlusion | 24% |
| Intracerebral hemorrhage | 10% |
| Stroke mimic | 27% |

Continuous variables are reported as mean (standard deviation). Ordinal variables are reported as median (interquartile range). Categorical variables are reported as proportions. NIHSS indicates the NIH stroke scale. If symptom onset was unwitnessed, time last known well was used as a surrogate.

As summarized in **Table 2**, the Openwater optical blood flow monitor demonstrated superior diagnostic performance relative to RACE and LAMS. Prehospital diagnostic performance was examined based on an estimated LVO prevalence of 5% and 10%. For 1,000 patient encounters, the Openwater optical blood flow monitor is expected to reduce both the number of false positives and false negatives as compared to RACE or LAMS (**Table 2**).

**Table 2.**
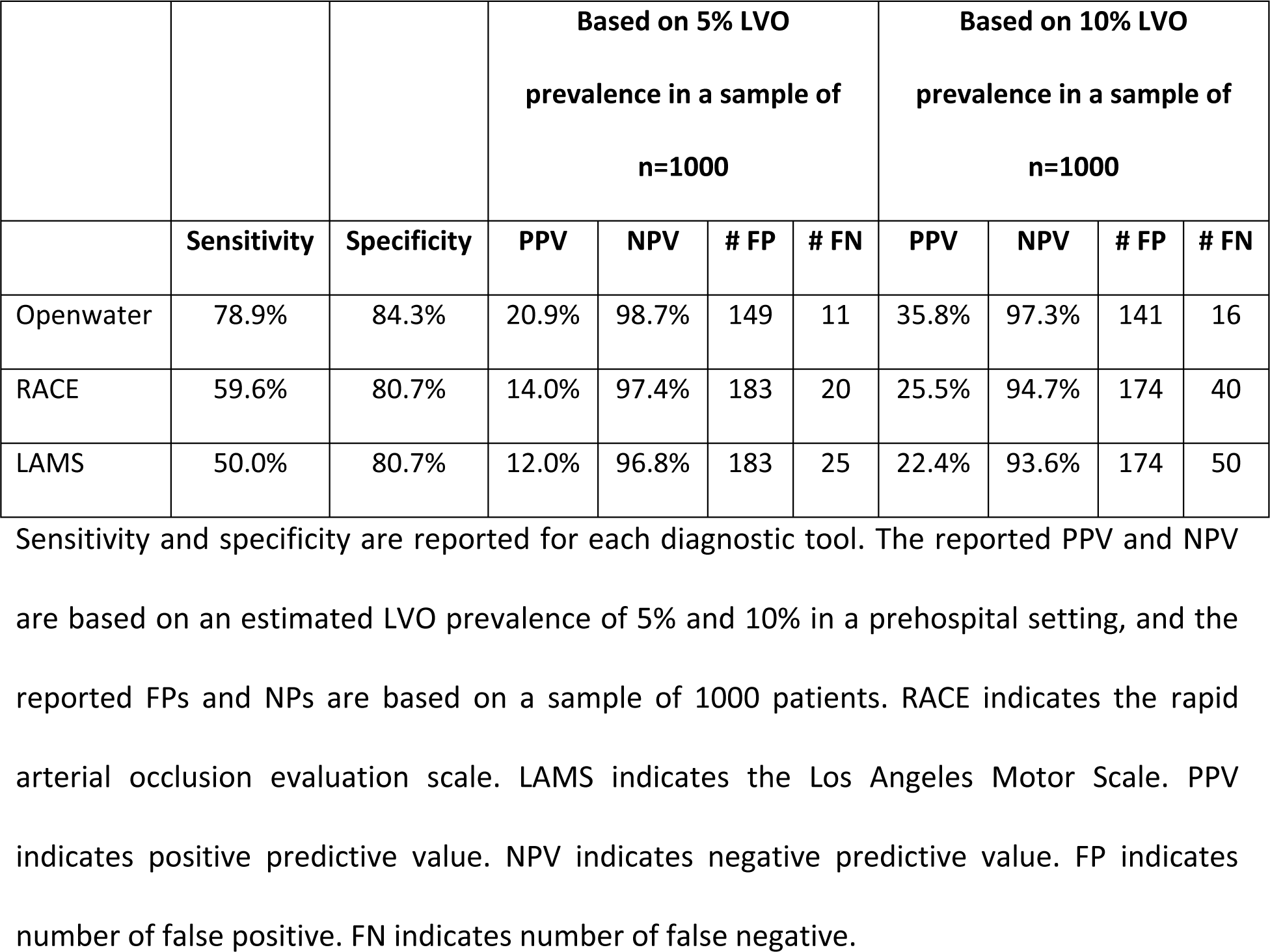
Diagnostic Performance.

Openwater’s AUROC was the largest when the full spectrum of data was considered (**Figure 3a**). **Figure S1** illustrates the relationship between Openwater performance and LVO cases using a logit function. For every one-unit increase in positive prediction score, the odds of LVO increased 18-fold (OR: 18.8, 95% CI: 7.52 - 47.14; p<.001). After applying the optimal Openwater threshold (Youden’s J) for LVO detection, the AUROC was higher than the clinically established RACE and LAMS thresholds (**Figure 3b**). **Figure S2** depicts the device performance for each of the five folds (*i.e.* used to facilitate the five-fold validation), and the Openwater performance was relatively consistent across all five models. Openwater performance was similar in patients with light and dark skin pigmentation (Fitzpatric scale 1-3 vs Fitzpatrick scale 4-6; **Figure S2**).

**Figure 3.**
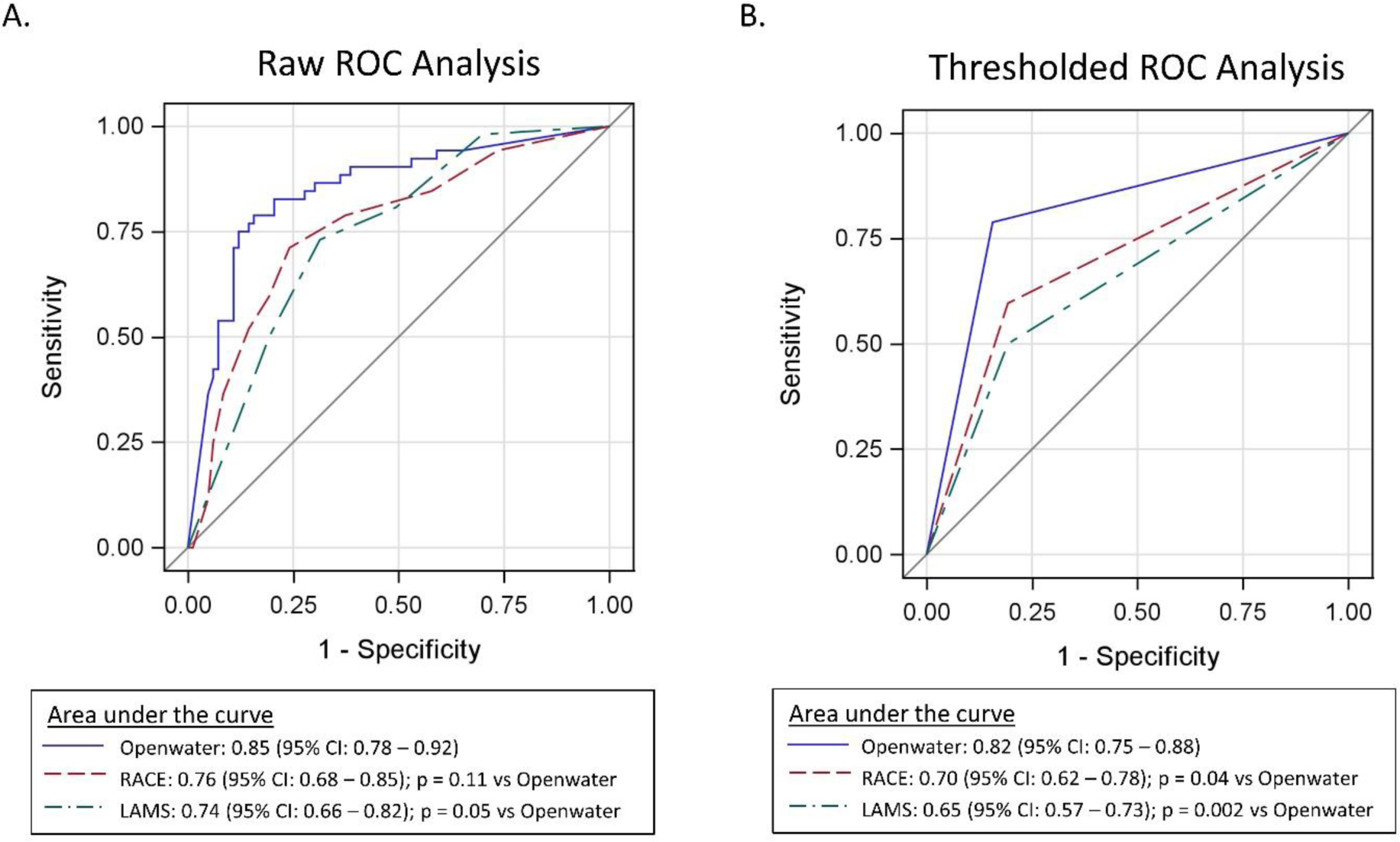
ROC analysis for LVO detection: (A) The receiver operator characteristic area under the curve is depicted when using raw scores. The area under the curve for the Openwater optical blood flow monitor is larger than that of LAMS. (B) The receiver operator characteristic area under the curve is depicted when using thresholded scores. The Openwater threshold was ≥ 0.80. The RACE threshold was ≥ 5. The LAMS threshold was ≥ 4. ROC indicates receiver operator characteristic. LVO indicates large vessel occlusion. RACE indicates the rapid arterial occlusion evaluation scale. LAMS indicates the Los Angeles Motor Scale.

## DISCUSSION

The Openwater optical blood flow monitor outperformed both RACE and LAMS for the detection of LVO in patients presenting for acute stroke evaluation. A clinically relevant increase in sensitivity was observed for the Openwater blood flow monitor without a cost to specificity, which ultimately yielded fewer false negatives and false positives. Reducing fast negatives is critical to early notification and routing of high likelihood LVO patients to thrombectomy capable or comprehensive stroke centers. Reducing false positives is similarly critical as it may reduce unnecessary patient routing and additional transport time. This capability is particularly relevant to non-LVO patients who are potentially eligible for intravenous thrombolysis. These encouraging results require validation in a prehospital cohort of potential with potential stroke.

The clinical relevance of false positives and false negatives is expected to vary geographically. For example, in urban environments, bypassing a primary stroke center may add minimal added travel time, whereas additional time may be expected in suburban and rural communities.^4,37^ Further, excessive bypassing may present a burden to comprehensive or thrombectomy-capable centers while leaving primary stroke centers underutilized. Future work may address the fact that the Openwater threshold can be titrated to emphasize either specificity or sensitivity to optimize care according to regional practices and logistics.^38^

Given the clinical demand, several techniques have been explored as potential prehospital LVO detectors.^16,17,39^ Some mobile stroke units are capable of performing CT angiography, which presents a tremendous opportunity to diagnose LVO in the field,^40^ but limitations, most notably cost, have hindered widespread implementation.^41^ Wearable or portable devices are appealing as they can be built into existing EMS infrastructure. Forest Devices developed a wearable cap that merges EEG and somatosensory evoked potentials (Forest Devices, Inc, Pittsburgh, PA). In an enriched cohort, similar to the current study, superior diagnostic accuracy was observed when compared with prehospital stroke scales.^19^ Although the EEG cap requires less than five minutes of set-up time, EEG may present additional logistical challenges in prehospital use. Cerebrotech developed a headset that leverages VIPS (Cerebrotech, Pleasanton, CA), a novel technology that uses low-power electromagnetic waves to detect asymmetry in bioimpedance, which in turn informs the likelihood of a large area of tissue injury.^20^ Though easy to use, the Cerebrotech device does not differentiate LVO from large hemorrhage or large ischemic stroke without LVO. In a single small study, transcranial ultrasound only detected 54% of LVOs.^42^ Although the combination of ultrasound and clinical exam improved diagnostic performance,^42^ ultrasound requires a degree of expertise that may not be reasonable to expect amongst EMS providers. Thorpe et al. used TCD to recognize CBF waveform features (quantified as the velocity curvature index and velocity asymmetry index) that achieved good diagnostic accuracy for LVO.^43^ Because the required technical expertise may limit broad implementation in the prehospital setting, Neural Analytics developed an autonomous system that obviates the need for an expert sonographer (NovaGuide™ TCD, Neural Analytics Inc, United States),^44^ but its potential role as in detecting LVOs in clinical practice remains unclear.

Biomedical optical techniques are particularly appealing in this context given the ability to directly probe microvascular hemodynamics in a portable and easy to use device. Cerebral oximetry based near-infrared spectroscopy (NIRS) is the most widely available optical technique and is often used as a surrogate of CBF,^45,46^ but changes in the NIRS signal may not mirror changes in CBF in the context of fluctuations in arterial oxygen saturation or cerebral metabolism,^47,48^ which may be particularly relevant limitation in acute stroke. Another optical technique, diffuse correlation spectroscopy (DCS), provides a direct measure of CBF by quantifying the speckle intensity fluctuations of near-infrared light that is scattered by tissues.^28,49^ Indeed, speckle fluctuations in space or time are the source of data for all emerging optical CBF methods. DCS has been used to monitor acute stroke physiology at the bedside,^30,31^ but signal-to-noise and acquisition frequency have to-date limited the ability of DCS to discern a high resolution CBF waveform for large source-detector separations.^50^ Speckle contrast optical spectroscopy (SCOS) uses a camera to measure speckle ensembles which in turn reflect changes in CBF, but again to-date the signal to noise ratio is not sufficient to resolve a high resolution CBF waveform.^51,52^ The Openwater optical blood flow monitor has some similarities to traditional SCOS but it also incorporates important methodological differences that allow it to overcome key limitations. In particular, short pulses of very intense laser light (rather than continuous light) permit signal-to-noise improvements and facilitate probing of tissue dynamics at very short time scales.^32^ The system also incorporates the cameras within the headset (instead of the conslole), which mitigates artifacts caused by cable motion. The ease of use and small portable design are also critical when considering the possibility of prehospital use. The Openwater system has previously been reported to resolve pulsatile CBF waveforms during the cardiac cycle, comparable to that of other high resolution instruments such as TCD.^32^

Despite the encouraging results, several limitations should be recognized. First, the optical scans were performed upon arrival in the Emergency Department rather than in the prehospital setting. By enrolling patients during the acute stroke evaluation, the cohort is reflective of the eventual target patient population, but the cohort was enriched with LVOs because of the large number of LVO transfers at the enrolling centers. Future work needs to evaluate prehospital feasibility and performance. Model performance may be limited by the relatively small sample size, but the five-fold validation offers added efficiency. The results are not reflective of a true test set, but similar performance across each fold provides some reassurance. In future work, a pre-specified model derived from these pilot results should be applied to an independent test cohort. The optical scan failed in 6% of cases as a result of poor headset contact which resulted in insufficient laser light and/or excess ambient light detected, and in 10% of cases due to excessive patient movement with resulted in failed pulse detection. Excluded patients had more severe strokes, and it is likely that patient movement and concomitant clinical care contributed to the scan failure in these cases. The brief data quality check performed at the beginning of each scan should be refined based on these observations to better alert the user of poor data quality and require an adjustment of the probes prior to data collection. These technical challenges need to be resolved before using the instrument in the prehospital environment where technical failures may be different or potentially more frequent. However, issues related to simultaneous clinical care (*i.e.* patients being moved or examined during the scan) may be less problematic in the prehospital setting where there are fewer providers. The diagnostic performance is unknown in small distal occlusions and may be clinically relevant to health care systems that routinely pursue EVT in such circumstances. The instrument probes the anterior circulation so is not expected to be sensitive to posterior circulation LVO, but no posterior circulation LVOs were enrolled in this study, so this may be directly explored in future work. There is also an opportunity to explore the subgroup of patients with mild clinical deficits in whom prehospital scales are particularly insensitive.^53^

## CONCLUSIONS

The Openwater optical blood flow monitor outperformed prehospital stroke scales for the detection of LVO in patients who presented to the Emergency Department for acute stroke evaluation. Future studies need to validate these findings in the prehospital environment in patients with suspected stroke. If validated, subsequent work can determine how to incorporate the results into routing workflow, and with further evaluation to clarify how the Openwater threshold can be titrated to meet the regional needs of different EMS and health care systems.

## Data Availability

The data and code that support the reported findings are available from the corresponding author upon reasonable request.

## Acknowledgements

The authors thank all study participants for the invaluable contribution and Peter Herzlinger for technical and engineering support.

